# Prediction Cause-Specific Mortality for Soft Tissue Cardiac Sarcoma Patients in the United States: A Competing Risk Analysis

**DOI:** 10.1101/2023.03.02.23286676

**Authors:** Roungu Ahmmad

## Abstract

**Objectives:** Several studies have revealed contradictory findings on survival by exposing cancer treatments with disparities of sociodemographic and tumor histologic factors. However, this study aimed to evaluate the effect of cancer treatments on mortality among patients with soft tissue cardiac sarcomas (CS) in the United States. We also estimated the overall survival probability through a competing risk nomogram for this notorious disease.

**Methods:** The data was taken from the US National Cancer Institute’s Surveillance Epidemiology, and End Results (SEER) -18 dataset, version 2020. Between the years 2000 and 2018. The study cohort included patients diagnosed with soft tissue cancer, including cardiac sarcoma. We computed the cumulative incidence function (CIF) and sub-distributional hazard by the Fine and Gray model for evaluating the risk of mortality. In addition, using a competing-risk nomogram we forecasted the overall survival probability for patients with soft tissue CS. Calibrations and Brier scores were used to validate and compare the prediction to ensure predictive accuracy.

**Results:** A total of 416 completed cases were selected for evaluation in this study. There was 66.5% mortality from soft tissue CS in the patients during this study period, while there was only 16.3% death from other diseases. For patients with soft tissue CS, the five-year cumulative incidence of cause-specific mortality was 74%, while only 18% was caused by other diseases. Non-whites, older age groups, and more advanced cancer stages all contributed to a higher cause-specific cumulative incidence, but sex was not a significant predictor of soft tissue CS deaths. Patients who underwent surgical intervention [sdHR: 0.55, 95%CI:0.28-0.98] and chemotherapy [sdHR: 0.29, 95%CI:0.02-0.36] on prime site had a significant decrease in CS death compared with no intervention, whereas primary systemic therapy and radiation intervention were not significantly decreased patients mortality. The patients who received surgery on the prime site survived 2, and 5 years above 70%, and 60%, respectively, whereas those who did not receive surgery on the prime site survived only 10%. Nomograms for assessing the hazard of mortality for patients with soft tissue cancers were well calibrated and had a good discriminatory ability.

**Conclusion:** Despite the small sample size, this study provided a reliable model-based prediction of the effect of cancer treatment on rare malignancies. The use of surgery and chemotherapy significantly reduced patient cause-specific mortality; however, the use of primary systemic treatment and radiation did not significantly reduce patient mortality among patients with soft tissue CS.

**Clinical Perspective:** 

**What is New?:**

- Patients with soft tissue Cardiac Sarcoma (CS) are more likely to die if they are older, at a distant stage, and have not received treatment.
- Among patients with CS, there is no significant association between race, gender, radiotherapy, or primary systemic therapy with mortality
- There is a greater reduction in mortality rate associated with chemotherapy and surgical intervention at the primary site than without these interventions.

**What are the Clinical Implications?:**

- A competing risk nomogram suggests that soft tissue CS is associated with a lower probability of cause-specific survival than other causes of mortality.
- The survival rate for patients who underwent surgery on prime sites was 70%, 55%, after two, and five years, whereas the survival rate for patients who did not undergo surgery was only 10%.
- A cause specific nomogram revealed that radiation intervention increases survivability compared to not receiving radiation intervention, and that age, sex, treatment, and cancer stages affected survival.

## BACKGROUND OF STUDY

Among the various types of sarcomas, soft-tissue sarcomas (STS) are malignant cell growths that may occur within connective tissue and other mesenchymal tissues (Demetri GD et al., 2010). A primary cancer of the heart is a cardiac sarcoma, which is a rare form of malignant (cancerous) tumor in the heart. Secondary cardiac tumors develop elsewhere in the body and spread to the heart (Tao, T.Y., et al., 2014). In most cases, cardiac sarcomas are angiosarcomas, which are most frequently found in the right atrium, blocking blood flow there (Li, X, et al., 2020) and patients usually identified of cardiac sarcoma only on post mortem analysis (Sarjeant, J.M., 2003).

Metastasizing soft tissue cardiac tumors occur 30 to 50 times more frequently than benign ones (Reynen, K, 1996), where one fourth of all heart tumors are malignant and are mostly cardiac sarcomas (Reynen, K, 1996; Silvermen N.A., 1980). Heart tumors occur rarely with an incidence of 0.001% to 0.03% in autopsy series, yet they are extremely deadly (Gutierrez JC, et al., 2007; Abraham KP, et al., 1990; Lam KY, et al., 1993). This disease does not exhibit any primary symptoms until it has developed metastatistic stages, which explains its high mortality rate (Silvermen, NA., 1980). Age is a common predictor of soft tissue CS, which affects those over 65 years old (Fine JP, et al., 1999). Approximately half of soft tissue CS patients die within a year of diagnosis, and their prognosis depends on the types and stages of cancer (Wolbers M., et al., 2009; Harrell F., 2015; Harrell FE, 2015). The incidence of metastatic heart cancer patients dying ranges from 1.5 to 13.9% with a general increase in mortality due to an increasing cancer incidence (Hanfling, SM., 1960, Anker, M.S., et al., 2020).

STS patients used to have a limited selection of treatment options. Patients with STS were treated, through chemotherapy, radiotherapy, and surgery at a main site or at a remote site (Miller KD, et al., 2018). A variety of different active ingredients are present in these treatments and many of them can lead to cancer of soft tissues, including cancer of the heart (Schöffski P, 2014). However, several recent studies have shown that combination chemotherapy provides optimal response rates and progression-free survival (Cella DF, et al., 1990; Fuller CD., et al., 2007; Wright CM, et al., 2020). Radiation therapy appears to play a significant role for survival in the incidence of soft tissue sarcomas (Loap P, et al., 2021; Parikh RC, et al., 2018), but the effect has not yet been determined for soft tissue CS very frequently. There is some evidence that patients with advanced tumor histology and different treatment combinations have varying survival, particularly for elderly patients. Even with advances in surgical and radiation techniques, patients with malignant pleura mausoleums have not had a better prognosis over the past four decades (Inoue, T., 2001). The treatment based on surgery provided advantageous in the earlier stages since cancer-directed surgery is independently associated with improved survival (Abdel-Rahman O., 2017; Taioli, E., et al., 2015; Shen W., et al., 2015) but very few study introduced for CS patients (Shen, W., et al., 2016). So, patients who have been diagnosed with regional or metastatic cardiac sarcoma may suffer from this illness upon diagnosis or later on after the illness recurs and the patient’s life expectancy drastically decreases (Dubal, PM, et al., 2016). The objectives of this study were to determine cancer stage-related survival and the impact of treatment by adjusting tumor histological and demographic factors.

## MATERIALS AND METHODS

In the United States, the Surveillance, Epidemiology and End Results Program (SEER) is a population-based cancer database covering over a quarter of the total population (Jemal A,, et al., 2004). The SEER program registries collect data about patient demographic characteristics, cancer types, stages, first treatment, and vital status at the time of final contact (Jemal A,, et al., 2004; Warren J, 2002; Suit HD., 1994). This study examined data from 18 registries cases from the Research Plus Database between the years 2000 and 2018, and collected data from 147 subsets of registries cases. According to our case study, there were 7.75 million cancer cases, and of these, 55 thousands of them were soft tissue cancers, which included heart cancers (Fig. 1). There were only 422 patients in the SEER database who were diagnosed with a very rare soft tissue sarcoma of the heart (CS) in the section 50.5. We were able to use the available statistics and treatment information related to rare diseases sarcomas in order to analyze cause-specific mortality and competing risk of death. The SEER data are available to the public, so an Institutional Review Board approval was not required, nor was SEER informed.

**Figure 1:**
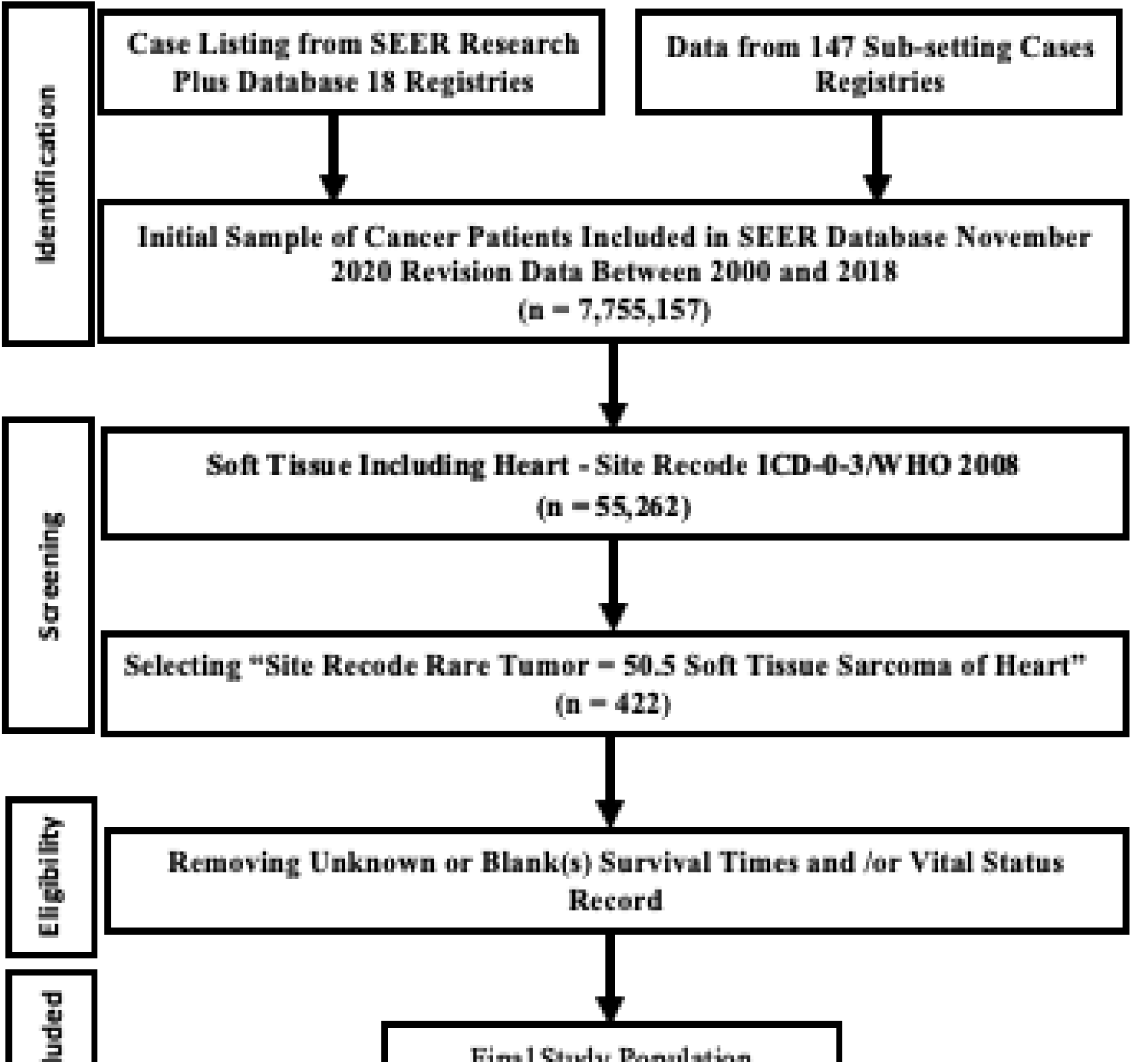
Flowchart of soft tissue including heart cancer identification in the SEER (18 registries) database. Data identification screening, eligibility and final included sample selection steps including the remove samples.

## PATIENTS CHARACTERISTICS AND VARIABLE SELECTION

The primary endpoints of the study were overall survival and competing mortality risks. In this study, patient survival was determined as the duration of time from diagnosis to death, while competing risk was determined as death due to cardiac sarcoma (cause-specific mortality) and death from all other-causes. Study subjects were followed from 2000 to 2018 and the following predictors were examined: cancer stage (localized, regional, distant), treatments (surgery at the primary site, chemotherapy, radiotherapy, and systemic treatment), tumor histology, and demographics (age, sex, race).

## METHODS

The statistical analysis was performed using R version 4.1.3 software, Stata/IC version 15.1 for Mac (64- Bit Intel) Revision 03 Feb 2020 Copyright 1985-2017 StataCorp LLC, Single-user Stata perpetual license, Serial Number 301506386406, Licensed to Md Roungu Ahmmad, The University of Mississippi Medical Center. The patients who received radiotherapy, chemotherapy, surgery on prime sites, or primary systemic therapy were monitored for cause-specific mortality and other causes of death. The median survival rate was determined by using Reverse Kaplan–Meier analysis. We summarized baseline characteristics and computed chi-square tests for categorical variables, and Kruskal-Wallis tests for continuous variables. We calculated the CIF and plotted the CIF curves for cause-specific and other-causes of mortality, respectively. The Fine-Gray proportional sub-distribution hazard model was used in order to estimate competing mortality risks (Fine JP, et al., 1999; Wolbers M., et al., 2009). This model meets the proportional hazard assumption by replacing the betas as a cosmetic modification. Further, the overall survival probability for patients who received cancer treatment can be predicted using a competing-risk nomogram. In order to validate the prediction accuracy, we used concordance (C) indices (Fine JP, et al., 1999), calibration plots (Wolbers M., et al., 2009) and Brier score. For analyzing competing mortality, building a model and nomogram, and evaluating the performance of the model, the R packages cmprsk, finalfit, pec, and rms were used. For both comparisons and inferences, p values were calculated using two-sided statistical testing and 5% significance level.

## RESULTS

### Baseline Characteristics

Table 1 presents the baseline characteristics of mortality grouped by cause of death. The study followed 245 individuals from 2000 to 2018, 227 died of cardiac sarcoma and 68 died from other causes; only 71 patients are censored. The median age was 48 (IQR: 35-62) years, 51% (n = 212) were female and 77.6% (n = 323) were white at the time of enrollment. Among the patients, 40 % (n = 149) had distant cancer stage, 29% (n = 106) had regional, and 86% (n = 357) had localized cancer stage (Table 1). A total of 85 (20.4%) patients took beam radiation therapy, and only 7 (1.2%) took beam with implants therapy (appendix table). Among all the patients, total 22% (n=92) received radiotherapy, 51% (n = 212) received chemotherapy, 54% (n=223) received surgery at the prime site, and 38% (n=117) treated the primary systemic therapy (Table 1).

**Table 1:** Overall mortality rate, five- person year cumulative incidence rate for cause specific mortality, and other causes of death for the patients of soft tissue cardiac sarcoma.

| Predictors | Overall,<br>N = 416<sup>1</sup> | Censored,<br>N = 71<sup>1</sup> | Death by CS,<br>N = 277<sup>1</sup> | Death by Other<br>Causes, N = 68<sup>1</sup> | p-value<sup>2</sup> |
| --- | --- | --- | --- | --- | --- |
| Age in Year | 48 (35, 62) | 43 (30, 56) | 48 (35, 60) | 58 (39, 73) | 0.001 |
| Sex |  |  |  |  | 0.9 |
| Female | 212 (51%) | 37 (52%) | 139 (50%) | 36 (53%) |  |
| Male | 204 (49%) | 34 (48%) | 138 (50%) | 32 (47%) |  |
| Race |  |  |  |  | 0.017 |
| Other (Non-White) | 93 (22%) | 14 (20%) | 72 (26%) | 7 (10%) |  |
| White | 323 (78%) | 57 (80%) | 205 (74%) | 61 (90%) |  |
| Cancer Stage |  |  |  |  | 0.085 |
| Localized | 106 (29%) | 20 (41%) | 71 (27%) | 15 (25%) |  |
| Regional | 114 (31%) | 18 (37%) | 77 (30%) | 19 (32%) |  |
| Distant | 149 (40%) | 11 (22%) | 113 (43%) | 25 (42%) |  |
| (Missing) | 47 | 22 | 16 | 9 |  |
| Surgery | 223 (54%) | 42 (59%) | 144 (52%) | 37 (55%) | 0.6 |
| (Missing) | 2 | 0 | 1 | 1 |  |
| Radiotherapy | 92 (22%) | 18 (25%) | 64 (23%) | 10 (15%) | 0.3 |
| Chemotherapy | 212 (51%) | 43 (61%) | 147 (53%) | 22 (32%) | 0.002 |
| Systemic Therapy | 117 (38%) | 38 (58%) | 69 (37%) | 10 (19%) | <0.001 |
| (Missing) | 109 | 5 | 90 | 14 |  |
| Total Tumor |  |  |  |  | <0.001 |
| One | 357 (86%) | 64 (90%) | 260 (94%) | 33 (49%) |  |
| Two | 48 (12%) | 6 (8.5%) | 14 (5.1%) | 28 (41%) |  |
| More than 2 | 11 (2.6%) | 1 (1.4%) | 3 (1.1%) | 7 (10%) |  |

<sup>1</sup> Median (IQR) or Non-Frequency (%)
<sup>2</sup> Kruskal-Wallis rank sum test; Pearson's Chi-squared test; Fisher's exact test

Figure 2 and Table 2 summarize the baseline characteristics and the cumulative incidence of cause-specific mortality and competing risk of death for the 5 person-year and 10 person-year time periods. The overall 5-person year mean cumulative incidence of death for cause-specific mortality was 74.8% (95% CI: 39.4–51.1%) and for other-causes of death was 14.3% (95% CI 6.1–11.2%) (table shown in appendix). Patients with older age, males, and non-white populations were more likely to experience a high cumulative incidence of cause-specific mortality. Patients with metastatic stages (distant) showed a higher mean cumulative incidence (5-person year: 74.0%, 10-person year: 78.8%) of cause-specific mortality compared to other causes (5-person year: 15.8%, 10-person year: 19.0%). Similarly, the incidence rate of distant stages found significantly higher than that of localized stages and regional stages. In comparison with patients who did not receive surgical intervention, those who underwent surgery at the primary site had a lower mean cumulative incidence for 5-years and 10-years (Table 2). The mean cumulative incidence of deaths among patients who received chemotherapy and radiotherapy was lower than among patients who did not receive treatment, however, primary systemic therapy had no effect.

**Figure 2:**
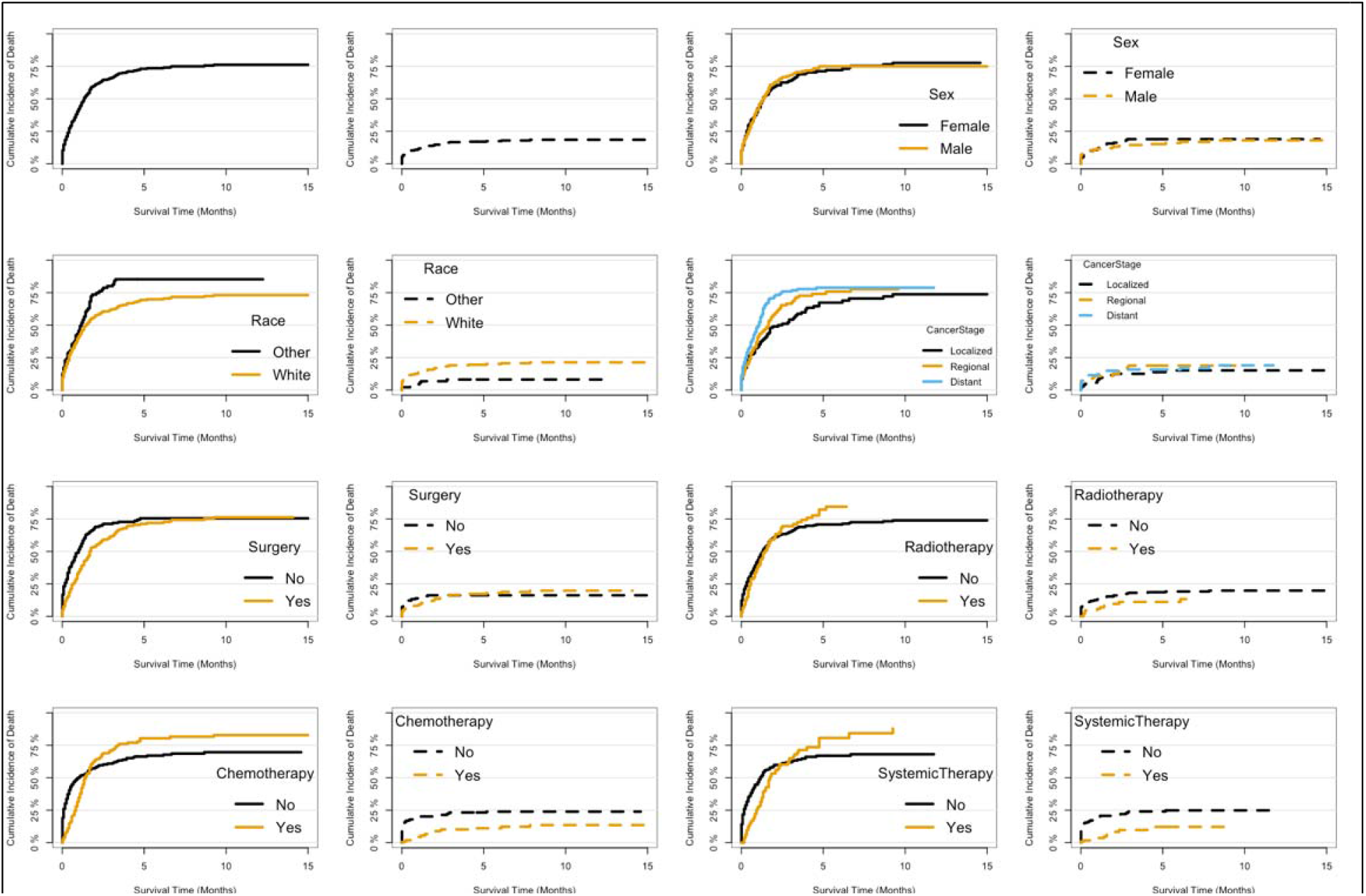
Cumulative incidence estimates of death according to the patient characteristics (solid line indicates cause-specific incidence rate; dotted line indicates other causes of death).

**Table 2:** Overall sample characteristics, 5-person year and 10-person year cumulative incidence rate for cause specific mortality and competing risk of death with Standard errors (SE) for patients with soft tissue cardiac sarcomas (N = 416):

| Predictors | Characteristics | Causes Specific Mortality |  |  |  | Competing Risk of Mortality |  |  |  |
| --- | --- | --- | --- | --- | --- | --- | --- | --- | --- |
|  |  | 5-Person<br>Year<br>Cumulative<br>Incidence | SE | 10-Person<br>Year<br>Cumulative<br>Incidence | SE | 5-Person<br>Year<br>Cumulative<br>Incidence | SE | 10-Person<br>Year<br>Cumulative<br>Incidence | SE |
| Age<50 Year | 231 (55%) | 0.562 | 0.001 | 0.754 | 0.0013 | 0.121 | 0.0008 | 0.181 | 0.002 |
| 51-70 Year | 127 (31%) | 0.738 | 0.002 | 0.807 | 0.0012 | 0.165 | 0.0007 | 0.139 | 0.011 |
| Above 70 Year | 59 (14%) | 0.764 | 0.001 | NA | NA | 0.370 | 0.0002 | NA | NA |
| Sex: Female | 212 (51%) | 0.712 | 0.003 | 0.777 | 0.0015 | 0.189 | 0.0004 | 0.189 | 0.003 |
| Male | 204 (49%) | 0.750 | 0.001 | 0.750 | 0.0011 | 0.151 | 0.0007 | 0.180 | 0.004 |
| Race: Non-White | 93 (22%) | 0.853 | 0.001 | 0.853 | 0.0011 | 0.081 | 0.0012 | 0.081 | 0.001 |
| White | 323 (78%) | 0.694 | 0.002 | 0.731 | 0.0013 | 0.195 | 0.0013 | 0.213 | 0.002 |
| Stages: Localized | 106 (29%) | 0.673 | 0.001 | 0.737 | 0.0015 | 0.138 | 0.003 | 0.151 | 0.001 |
| Regional | 114 (31%) | 0.740 | 0.001 | NA | NA | 0.188 | 0.0007 | NA | NA |
| Distant | 149 (40%) | 0.740 | 0.002 | 0.788 | 0.0014 | 0.158 | 0.0006 | 0.190 | 0.012 |
| Surgery: No |  | 0.755 | 0.001 | 0.750 | 0.0013 | 0.162 | 0.0012 | 0.162 | 0.003 |
| Yes | 223 (54%) | 0.712 | 0.002 | 0.763 | 0.0013 | 0.173 | 0.0008 | 0.198 | 0.001 |
| Chemotherapy: No |  | 0.804 | 0.003 | 0.828 | 0.0011 | 0.232 | 0.0008 | 0.239 | 0.003 |
| Yes | 212 (51%) | 0.664 | 0.002 | 0.697 | 0.0012 | 0.111 | 0.0005 | 0.135 | 0.002 |
| Radiotherapy: No |  | 0.822 | 0.001 | 0.739 | 0.0011 | 0.186 | 0.0006 | 0.198 | 0.005 |
| Yes | 212 (51%) | 0.707 | 0.001 | NA | NA | 0.111 | 0.0005 | NA | NA |
| Syst. Therapy: No |  | 0.661 | 0.002 | 0.681 | 0.0013 | 0.249 | 0.0002 | 0.249 | 0.002 |
| Yes | 117 (38%) | 0.806 | 0.003 | NA | NA | 0.122 | 0.0002 | NA | NA |

Patients who did not undergo surgery on the prime sites experienced a significantly higher rate of death (5-person year: 75.5% %, 10-person year: 75%) compared with other-causes of mortality (16.2 %, 16%). Radiotherapy and chemotherapy did not lead to a different mortality rate for cause-specific cancers (see appendix table). In contrast, patients who didn’t receive radiotherapy or chemotherapy had a high cumulative incidence of other-causes of death. According to the study, radiation therapy and chemotherapy are not beneficial for patients with CS, while surgery can reduce mortality (Table shown in appendix).

Five-year estimates of the crude cumulative incidence of cause-specific death and other causes of death by age at diagnosis, tumor size, sex, race, stage, and treatment are presented in Table 2. According to Fig. 2 (plot 1 and 2), patients who have a high cumulative incidence of cause-specific mortality compare with other causes of death have been identified. Among patients with soft tissue including heart cancer, the 6- year overall cumulative incidence rate of death was 74% on the other hand, only 18% cumulative incidence (CI) for other cause of mortality. The CI of cause-specific mortality was higher in individuals with characteristics of older age, male and non-white population (fig. 2). Comparing distant cancer stages to localized and regional stages, the patient with distant cancer stage predicted higher cumulative incidence of death for CS patients but did not predict cumulative incidence of death for other causes.

Surgery resulted in decreased probability of cause-specific death. Radiation therapy had not significantly affect for reduction the mean cumulative incidence rate for CS patients but for other causes, approximately 10% reduced the mean cumulative incidence of death for the patients with soft tissue including heart cancer (Fig2). Similar finding found for chemotherapy and systemic therapy. Among patients with soft tissue including heart cancer, the 10-year overall survival, mean cumulative incidence of death was significantly lower for those who had received surgery on the prime site, compare with those who had not received surgery for the patients with soft tissue CS. Therefore, the exposure group of patients who were treated with systemic therapy and surgery on the prime site had a significantly reduced risk of cause-specific mortality at most 10 years, but radiotherapy and chemotherapy had no significant effects on patient mortality. In the same way, chemotherapy significantly reduced the likelihood of cumulative incidence in patients with other causes.

Table 3 and Figure 3B present the coefficients and sub-distribution hazards ratios (sdHRs) based on the competing-risk model for event-free survival, cause-specific death, and other causes of mortality of soft tissue CS. Figure 3A illustrates the all-cause mortality hazard plots with 95% confidence intervals and level of significance. In this study, age, gender, race, and stage of cancer were adjusted. Surgery on the primary site and chemotherapy significantly reduced all-cause mortality but primary systemic therapy and radiation intervention had no significant effect on soft tissue CS.

**Table 3:** Proportional Sub distributional hazard model for cause specific mortality and competing risk of death for other causes for the patients with soft tissue cardiac sarcomas.

| Predictors | Competing Risk Model |  |  | Cox Model |
| --- | --- | --- | --- | --- |
|  | Event-Free Survival<br>HR [95% CI] | Other Cause of Mortality<br>HR [95% CI] | Cause-Specific Hazard Rate<br>HR [95% CI] | All Cause Mortality<br>HR [95% CI] |
| Age: Below 50 Years | 1 | 1 | 1 | 1 |
| 51-70 Years | 1.432 [1.03 - 1.99] | 1.389 [0.96 - 2.00] | 1.800 [0.85 - 3.81] | 1.399 [1.02 - 1.96] |
| Above 70 Years | 2.337 [1.50 - 3.64] | 1.541 [0.89 - 2.68] | 6.012 [2.67 - 13.53] | 2.189 [1.47 - 3.56] |
| Sex: Female | 1 | 1 | 1 | 1 |
| Male | 0.966 [0.73 - 1.28] | 0.966 [0.71 - 1.32] | 0.848 [0.45 - 1.60] | 0.994 [0.75 - 1.32] |
| Race: Non-White | 1 | 1 | 1 | 1 |
| White | 0.832 [0.59 - 1.17] | 0.716 [0.50 - 1.03] | 1.986 [0.79 - 4.98] | 0.848 [0.59 - 1.16] |
| Cancer Stage: Localized | 1 | 1 | 1 | 1 |
| Regional | 1.432 [0.97 - 2.11] | 1.264 [0.82 - 1.95] | 2.204 [0.97 - 5.00] | 1.442 [0.98 - 2.12] |
| Distant | 2.080 [1.44 - 3.01] | 1.963 [1.31 - 2.95] | 2.399 [1.04 - 5.51] | 2.017 [1.43 - 2.99] |
| Surgery: No | 1 | 1 | 1 | 1 |
| Yes | 0.613 [0.44 - 0.86] | 0.649 [0.44 - 0.95] | 0.550 [0.28 - 0.98] | 0.605 [0.43 - 0.84] |
| Radiotherapy: No | 1 | 1 | 1 | 1 |
| Yes | 0.794 [0.56 - 1.12] | 0.887 [0.61 - 1.28] | 0.482 [0.18 - 1.28] | 0.824 [0.58 - 1.16] |
| Chemotherapy: No | 1 | 1 | 1 | 1 |
| Yes | 0.455 [0.30 - 0.70] | 0.645 [0.40 - 1.03] | 0.290 [0.02 - 0.36] | 0.448 [0.29 - 0.69] |
| Systemic Therapy: No | 1 | 1 | 1 | 1 |
| Yes | 1.235 [0.80 - 1.91] | 1.149 [0.72 - 1.84] | 2.335 [0.55 - 9.98] | 1.274 [0.83 - 1.98] |
| Model Performance |  |  |  |  |
| Concordance | 0.705 (se = 0.02) | 0.695 (se = 0.024) | 0.843 (se = 0.031) | 0.706 (se = 0.02) |

**Figure 3:**
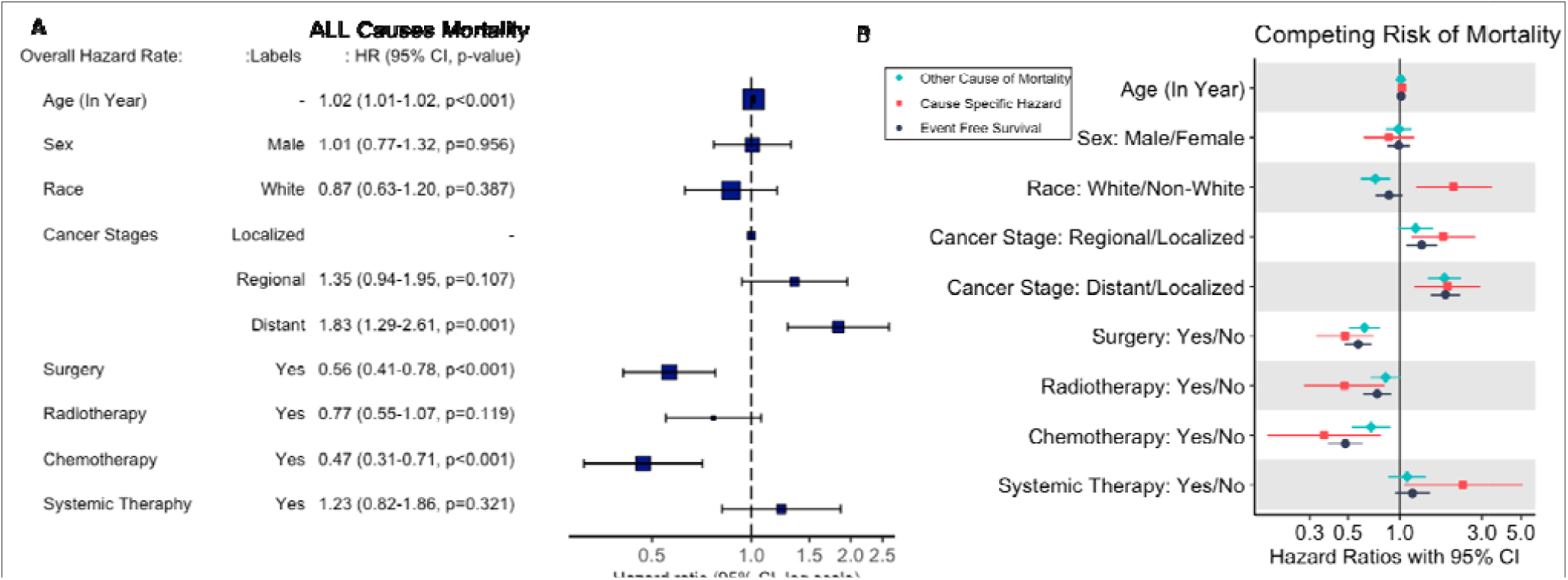
Overall hazard rate for all-cause mortality and effect of cancer treatments by adjusting demographic factors and tumor histological factors (Concordance: 0.706 (se = 0.02))

In the competing risk model, patients with distant cancer stage were more likely to die of their disease than patients with localized and regional stages, with an SDHR for localized cancer stage of 2.2 (95% CI: 0.97-5.00) and distant cancer stage of 2.4 (95% CI: 1.04-5.51). Moreover, patients undergoing surgery at prime sites experienced a significantly reduced risk of CS death compared with those not undergoing surgery, with a sdHR of 0.550 [0.28 - 0.98]. Chemotherapy significantly decreased cause-specific mortality (sdHR: 0.290, 95%CI: 0.02 to 0.36), however, primary systemic therapy and radiation intervention did not significantly affect patients’ survival. For comparison, we examined the impact of cancer treatment on survival with event-free survival and other-causes of death, where similar results were noted.

According to Fine and Gray’s model, Figure 4 represents the competing risk nomogram predicting the probability of cause-specific death for patients with soft tissue CS. In the nomogram, the characteristics of the patient are located on the variable row, and a vertical line is drawn straight up to the points row to assign a value. For example Figure 3A, if a vertical line is drawn directly above the point row, we obtain a score of 30 points for an 30-year-old patient. Then, continue this process for each subsequent row till all the variables have been accumulated. The probability of cause-specific survivability may be calculated by adding up the total points and drawing a vertical line from the row of total points. If the sum of points for each variable is 90, then for patients with CS at age 30 at the end of two years, the survival probability would be approximately 48%, and after five years, it would be approximately 32% (Figure 4A).

**Figure 4:**
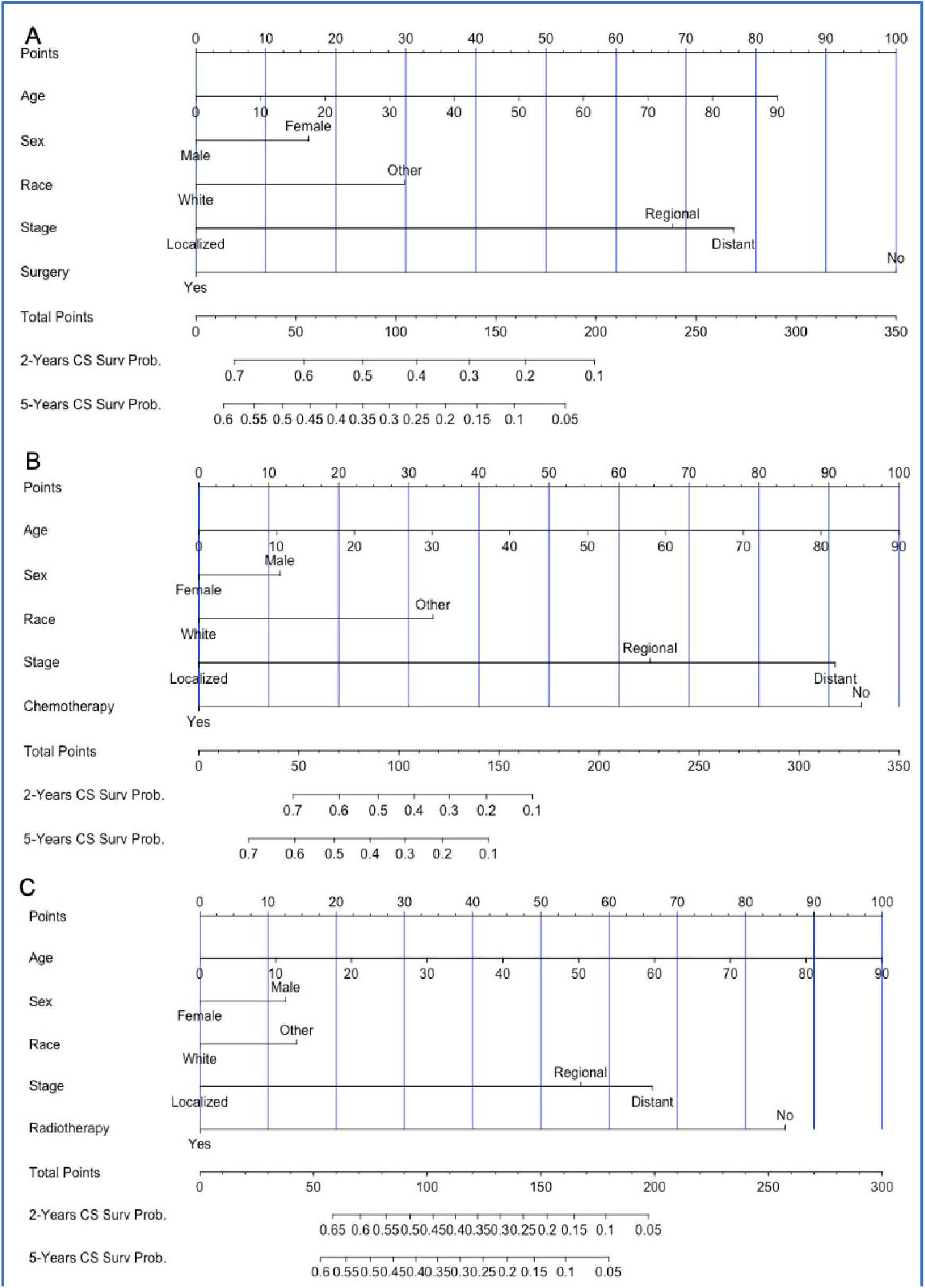
Nomogram for predicting 2-, 5- year probabilities of cause specific mortality in patients with soft tissue cardiac sarcomas according to plot 1 shown there is no interaction between predictors and plot 2 shown that the interaction between age and sex and interaction between surgery and cancer stages.

Furthermore, patients receiving only surgical intervention on the prime site would be likely to live for 2 and 5 years with a survival probability above 70%, and 60%, respectively, while patients not receiving surgical intervention would have survival probabilities below 10%. Similarly, patients who only receive chemotherapy have a survival probability above 70%, while patients without chemotherapy have survival probabilities below 10%. A patient with CS who only received radiation intervention has a survival probability of more than 70% for 2 and 5 years, while a patient without radiotherapy has a survival probability below 10% (Figure 4C)

In Figure 5, we present the time-dependent AUC and prediction error plot. Nomogram 1 includes only patients who have received only surgical treatment. By adjusting the baseline predictors, we fitted a competing risk model and measured the overall probability of survival. Similarly, for nomograms 2 and 3 we filtered the patients who received only chemotherapy and radiation treatments and fitted competing risk models. To make comparisons, we also consider the null model where no cancer treatments are given. On the basis of these results we computed the area under the ROC curve (AUC) and Brier Score (prediction error) for all our models. According to the same model structure, all the nomograms were well calibrated with higher time dependent AUC than the null model (Figure 5). Furthermore, the time dependent Brier scores were lower than the null model, which suggests good model reliability and higher prediction capability.

**Figure 5:**
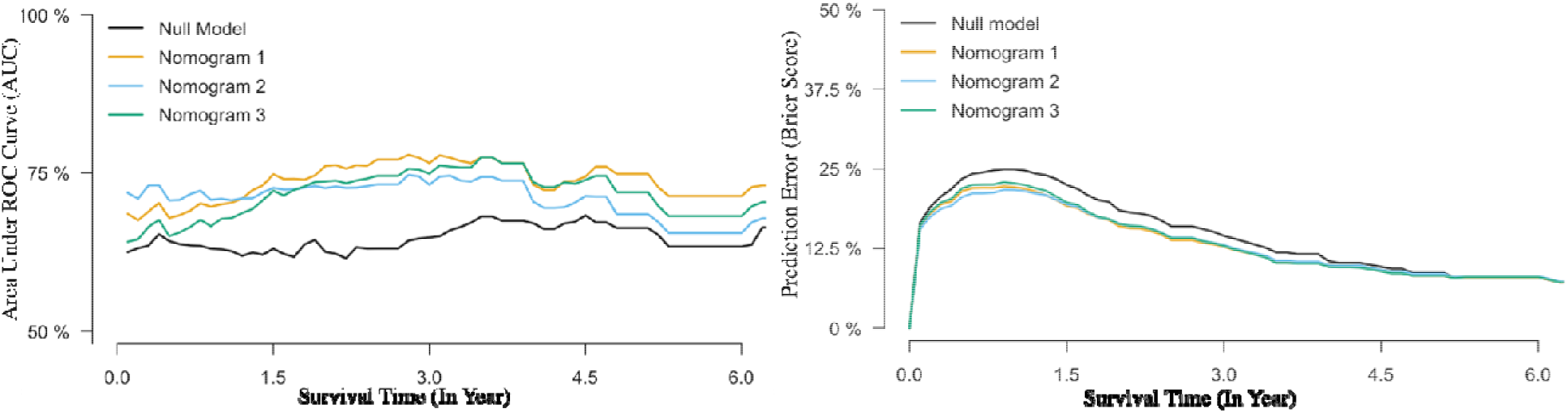
Time-dependent Area Under the ROC curve (Concordance Index) and Prediction Errors for the competing risk nomograms 1 The AUC presented for the three risk nomograms and compared with the Null model (Without covariates) 2 Brier Score (Prediction errors)

## DISCUSSION

The study assessed the probability of death among patients with soft tissue CS diagnosed between 2000 and 2018 based on the SEER dataset. Among 416 complete cases, 66.5% died from soft tissue CS, while only 16.3% died from other conditions. Patients with soft tissue CS had a five-year cumulative cause-specific mortality rate of 74%, while only 18% of deaths were caused by other diseases. There was a higher cause-specific cumulative incidence for non-whites, older age groups, and advanced stages of cancer; however, sex did not seem to be a significant predictor of soft tissue CS death. In addition, using a competing-risk nomogram, we projected the overall survival probability of patients with soft tissue cancer. It is the first study that has attempted to evaluate the risk of mortality for patients with CS associated with cancer treatments.

The results of the study did not reveal any difference between men and women in terms of death due to CS. Furthermore, we fitted the traditional Cox proportional hazard model for overall mortality outcomes for these patients and found that sex was not a significant predictor of mortality for patients with soft tissue CS. According to other studies, mortality rates did not significantly differ between the sexes among patients who had head and neck adenoid cystic carcinoma, basaloid squamous cell carcinoma (Shen, et al., 2018, Shen, et al., 2017), whereas a study in the SEER registry reported a better survival outcome among women diagnosed with head and neck cancer from 1973–2007 (Ellington, et al., 2012). Patients’ age is an important factor that predicts their chance of survival. We predicted overall survival probability for continuous 10 years increment in competing risk nomogram. Age was found to be a significant determinant of cause-specific mortality and other causes of death. We found that, according to the competing risk nomogram, patients aged 30 years have a survival probability of 48 % and 30 % respectively when they have soft tissue CS, while those aged 50 years have a survival probability of around 20 %. In accordance with other studies (Shen et al., 2018, Shen et al., 2017, Ellington et al., 2012) older patients were more likely to die of CS. Similarly, to our findings, the competing risk of mortality significantly increased with the patient’s age, which was in agreement with the results of our study. Similarly, white patients have significantly higher survival probability compare with non-white patients.

A high percentage of cause-specific deaths tends to occur among patients who were at a distant stage. Among 416 patients, 40% (n = 149) were in metastasis stages, where 113 (43%) death by soft tissue CS and around 29% and 31% of the patients were in localized and regional stages. 5-person years cause specific cumulative incidence for distant stages have significantly higher compare with localized and regional stages, but other cause of death was not significantly differ. In distant stage, the patient had 2- fold higher risk of all cause mortality compare with localized stage whereas cause specific death rate was 2.4 times higher. From competing risk nomogram, the patient who received surgery in localized and regional stages, have around 30%-40% increased the survival for 2 years. However, in distant stage, approximately 15% change the patient survive at least survival 2 years. Additionally, the patient underwent radiation intervention on the prime site, the survival probability did not increase a lot in all cancer stages, however, chemotherapy increase survival for patient with soft tissue CS. The patients who received chemotherapy in regional stage, at least 35% increased survival for 2 years and approximately 35% increased for 6 years. The patient in distant stages, chemotherapy significantly is more effective for patients’ survival compared with surgery and radiation intervention. Some studies shown sugary on squamous cell carcinoma, head and neck adenoid cystic carcinoma increased patients’ survival but radiotherapy was not significant effect on patient survival studies (Shen et al., 2018, Shen et al., 2017, Ellington et al., 2012). Another study shown surgery and radiotherapy on basaloid squamous cell carcinoma of head and neck increased patients’ survival (Shen, W., 2017). Consistent with these, our study found that, chemotherapy and surgery on the prime site for patient with soft tissue CS reduced mortality significantly on the other hand, systemic therapy and radiation intervention did not improve patient survival. Compare with all-cause mortality, these outcomes are more reliable and is consistent with other studies that used the SEER cohort (Lam KY., 1993; Dubal, PM., et al., 2016). Since radiotherapy is usually offered to those who cannot undergo surgery, some people who receive radiotherapy may have the most advanced disease. It may be because of this that radiotherapy didn’t improve the prognosis in patients with soft tissue cancer (Abraham, KP., et al., 1990). In the presence of chemotherapy, CIF is significantly decreased for the patients with soft tissue CS compared to that without those treatments and adjusting with other predictors, chemotherapy significantly decreased mortality. For other cause of mortality, event free survival and all-cause mortality, chemotherapy significantly improved patient survival. There have been some studies conducted on soft tissue sarcomas to predict survival based on the Cox model (Wolbers, M., et al., 2009; Harrell F., 2015; Miller KD., et al., 2018). This study confirmed the findings of those studies and provided additional inferences about the effects of treatments on soft tissue CS.

Our study differed from previous studies due to the endpoint of the study and the methodology used to model and construct the nomograms. In a few publications, competing-risk models have been argued, including sarcoma, breast, prostate, melanoma, thyroid, and head and neck squamous cell carcinoma (Cella, DF., et al., 1990; Fuller CD., et al., 2007; Taioli, E., et al., 2015), this study is the first to develop a nomogram-based model of survival prediction in soft tissue cancer and investigate the effect of cancer treatments. Another strength of our study can be attributed to the simplicity of the model and nomogram, which are easily understandable by clinicians and are more comparable.

## LIMITATIONS

While we were able to use a large dataset from the SEER, our study had some limitations. Due to the lack of public access to SEER, some clinicopathological factors (such as surgical margins, perineural invasion, solid tumors, and p53 positivity) were not included in this study. Furthermore, although histopathological details have proven to be critically important in predicting survival, we cannot use them in the model because histological subtypes of CS are not included in the SEER public-use dataset. Additionally, SEER does not provide data on cancer recurrences. A relatively short follow-up is another limitation of this study, as we have only a limited number of patients available during the study period. A longer follow-up may improve the nomogram and increase the model’s accuracy, especially for patients with soft tissue CS. The nomogram and model performance might be effective, however external validation with other cohorts is still needed to confirm its efficacy.

## CONCLUSIONS

Our study calculated the cumulative incidence of cause specific mortality and other causes of death for patients with soft tissue cardiac sarcoma using a large, population-based dataset. We evaluated competing risk mortality, event-free survival for patient with soft tissue CS and built nomograms using variables that could be obtained easily and modeled the probability of cause-specific death utilizing competing risk model. Surgery and chemotherapy significantly improved patients survival however, primary systemic therapy and radiation intervention did not significantly reduced mortality for patient with soft tissue CS.

## Data Availability

All data produced in the present study are available upon reasonable request to the authors

https://seer.cancer.gov/about/

## ACKNOWLEDGEMENT

The authors would like to thank the NCI for open access to their SEER database. The opinions or views expressed in this paper are those of the authors and do not represent the opinions or recommendations of the NCI.

## Appendex

**App Table:** Summary statistics grouped by Cause specific deatha and other casues of death with percentages and p values.

| Characteristics | Total N (%) | Couuse-specific Death |  |  | Death from other Causes |  |  |
| --- | --- | --- | --- | --- | --- | --- | --- |
|  |  | Survivors N (%) | Dead N (%) | pval | Survivors N (%) | Dead N (%) | pval |
| Survival Months: Median (IQR) | 9.0 (2.0 to 20.0) | 10.0 (1.0 to 27.5) | 9.0 (2.0 to 18.0) | 0.247 | 10.0 (2.8 to 21.0) | 2.5 (0.0 to 14.8) | 0.001 |
| Age: Median (IQR) | 48.0 (35.0 to 62.0) | 50.0 (32.5 to 64.5) | 48.0 (35.0 to 60.0) | 0.759 | 47.0 (35.0 to 60.0) | 57.5 (39.2 to 73.0) | 0.003 |
| Age Group: <30 Years | 77 (18.5) | 32 (23.0) | 45 (16.2) | 0.042 | 64 (18.4) | 13 (19.1) | <0.001 |
| 30-40 Years | 65 (15.6) | 19 (13.7) | 46 (16.6) |  | 61 (17.5) | 4 (5.9) |  |
| 41-50 Years | 89 (21.4) | 21 (15.1) | 68 (24.5) |  | 79 (22.7) | 10 (14.7) |  |
| 51-60 Years | 76 (18.3) | 23 (16.5) | 53 (19.1) |  | 66 (19.0) | 10 (14.7) |  |
| 61-70 Years | 51 (12.3) | 17 (12.2) | 34 (12.3) |  | 41 (11.8) | 10 (14.7) |  |
| >70 Years | 58 (13.9) | 27 (19.4) | 31 (11.2) |  | 37 (10.6) | 21 (30.9) |  |
| Sex: Female | 212 (51.0) | 73 (52.5) | 139 (50.2) | 0.729 | 176 (50.6) | 36 (52.9) | 0.822 |
| Male | 204 (49.0) | 66 (47.5) | 138 (49.8) |  | 172 (49.4) | 32 (47.1) |  |
| Race: Non-White | 93 (22.4) | 21 (15.1) | 72 (26.0) | 0.017 | 86 (24.7) | 7 (10.3) | 0.014 |
| White | 323 (77.6) | 118 (84.9) | 205 (74.0) |  | 262 (75.3) | 61 (89.7) |  |
| Cancer Stage: Distant | 149 (35.8) | 36 (25.9) | 113 (40.8) | 0.206 | 124 (35.6) | 25 (36.8) | 0.829 |
| Localized | 106 (25.5) | 35 (25.2) | 71 (25.6) |  | 91 (26.1) | 15 (22.1) |  |
| Regional | 114 (27.4) | 37 (26.6) | 77 (27.8) |  | 95 (27.3) | 19 (27.9) |  |
| (Missing) | 47 (11.3) | 31 (22.3) | 16 (5.8) |  | 38 (10.9) | 9 (13.2) |  |
| Radiation Type: None | 324 (77.9) | 111 (79.9) | 213 (76.9) | 0.785 | 266 (76.4) | 58 (85.3) | 0.265 |
| Beam | 85 (20.4) | 26 (18.7) | 59 (21.3) |  | 76 (21.8) | 9 (13.2) |  |
| Beam with Implants | 7 (1.7) | 2 (1.4) | 5 (1.8) |  | 6 (1.7) | 1 (1.5) |  |
| Radiotherapy: Yes | 92 (22.1) | 28 (20.1) | 64 (23.1) | 0.575 | 82 (23.6) | 10 (14.7) | 0.147 |
| No | 324 (77.9) | 111 (79.9) | 213 (76.9) |  | 266 (76.4) | 58 (85.3) |  |
| Chemotherapy: Yes | 212 (51.0) | 65 (46.8) | 147 (53.1) | 0.267 | 190 (54.6) | 22 (32.4) | 0.001 |
| No | 204 (49.0) | 74 (53.2) | 130 (46.9) |  | 158 (45.4) | 46 (67.6) |  |
| Systemic Therapy: Yes | 117 (28.1) | 48 (34.5) | 69 (24.9) | 0.67 | 107 (30.7) | 10 (14.7) | 0.002 |
| No | 190 (45.7) | 72 (51.8) | 118 (42.6) |  | 146 (42.0) | 44 (64.7) |  |
| (Missing) | 109 (26.2) | 19 (13.7) | 90 (32.5) |  | 95 (27.3) | 14 (20.6) |  |
| Surgery Prime site: Yes | 223 (53.6) | 79 (56.8) | 144 (52.0) | 0.384 | 186 (53.4) | 37 (54.4) | 0.912 |
| No | 191 (45.9) | 59 (42.4) | 132 (47.7) |  | 161 (46.3) | 30 (44.1) |  |
| (Missing) | 2 (0.5) | 1 (0.7) | 1 (0.4) |  | 1 (0.3) | 1 (1.5) |  |
| Total Tumor Prime site: One | 357 (85.8) | 97 (69.8) | 260 (93.9) | <0.001 | 324 (93.1) | 33 (48.5) | <0.001 |
| Two | 48 (11.5) | 34 (24.5) | 14 (5.1) |  | 20 (5.7) | 28 (41.2) |  |
| More than Two | 11 (2.6) | 8 (5.8) | 3 (1.1) |  | 4 (1.1) | 7 (10.3) |  |

